# Exponential distribution of large excess death rates in Europe during the COVID-19 outbreak in the spring of 2020

**DOI:** 10.1101/2020.09.20.20198283

**Authors:** José María Martín-Olalla

**Author notes:** Twitter: @MartinOlalla JM.

## Abstract

Excess death rates *E* during the spring of 2020 are computed in *N* = 540 level 3 European territorial units for statistics —NUTS3 in Belgium (40), France (96), Italy (110), Netherlands (44), Spain (50), Sweden(21) and United Kingdom (179)— from 2020 provisional week deaths, the population numbers for 2020, and observations in previous years (reference or baseline), all of them obtained from Eurostat web page.

Excess death rates are classified in three tiers. Largest 27 excess death rates (tier 1, *E >* 1721*×* 10^−6^) were distributed exponentially with empirical complementary cumulative distribution function (empirical survival function) *S* following *S* ∝ 2^−*E/ε*^ with *ε*_1_ = 958(42) *×* 10^−6^. Tier 2 (the next 52 largest excess death rates, *E* < 1142 *×* 10^−6^ also distributed exponentially with *ε*_2_ = 379.5(89) *×*10^−6^. Tier 3 (smallest 460 excess death rates) were distributed normally.

The results suggests that when, within some regions, the outbreak is above a threshold, interaction with neighbouring region become less relevant and the outcomes —excess death rates— become exponentially distributed as it happens with independent events.

## I. Introduction

Excess death rates[1] relative to a consistent reference value based on historical records are a key quantity to understand the impact of a pandemic like the one associated to the illness designated COVID–19 which is caused by the severe acute respiratory syndrome coronavirus 2 (SARS-CoV-2)[2]. The illness caught worldwide attention since its identification in late 2019 and impacted largely in several countries of Europe during the spring of the year 2020.[3] Mortality monitors like EuroMoMo recorded extremely large anomalies in Italy, Spain, and United Kingdom among others. Societal responses in the form of nationwide lock-downs were taken by governments at the spring of 2020.

Many empirical quantities shows a normal (Gaussian) distribution. The sample average value and the sample standard deviation suffices to characterize the distribution as the density of probability quickly falls to zero which increasing *x* (*∝* exp(−*x*^*2*^)). Many other societal quantities are characterized by the presence of abnormally large events[4]. A nice example of this is the distribution of population in municipalities from villages to mega cities. Likewise the heavy tail distribution for the risk of contagious diseases is also an issue of recent interest.[5, 6]

This work will analyze the distribution of excess crude deaths in European level 3 territorial units for statistics in Belgium, France, Italy, Netherlands, Spain, Sweden, and United Kingdom. Helped by population numbers, excess death rates *E* will be computed for the spring of 2020. The work aims to understand the distribution of events along the 540 territorial units that entered in the analysis with focus in characterizing abnormally large excess deaths. Notice that abnormally low (negative) excess deaths are less likely to occur since mortality in modern societies comes mainly from natural causes. However societal responses in the form of lock-downs may favor mild negative excess deaths.

## II. DATA SET

Historical records of mortality in Europe can be obtained from “Weekly deaths–special data collection”^1^ disaggregated by the Nomenclature of Territorial Units for Statistics (NUTS). A visual inspection of the excess of deaths in 2020 relative to 2019 (see Supplementary Material figure 1) find signficant anomalies during the spring in Belgium, France, Italy, Netherlands, Spain, Sweden, and United Kingdom. For the forthcoming analysis weekly deaths for level 3 of NUTS —the smallest available unit— will be analyzed in these countries were considered. French overseas departments (code FRY) and Canary Islands (code ES7) did not enter in the analysis. Population numbers were retrieved from Eurostat “Population on 1 January by age group, sex, and NUTS 3 region”^2^ table which lists values from 2019 to 2014. Population values for 2020 were obtained after rescaling 2019 regional population numbers with nationwide 2020 population obtained from table `tps00001`^3^. For Spain numbers were taken from table `31304` at the National Statistics Institute^4^ which includes numbers from 2001 until 2020. Weekly population values were obtained for every ISO-8601 week by linear interpolation from January 1, 2020 backwards. Population in 2020 was set constant irrespective of week number. As of 2020 the total population in the seven countries is 278 746 738.

**FIG. 1.**
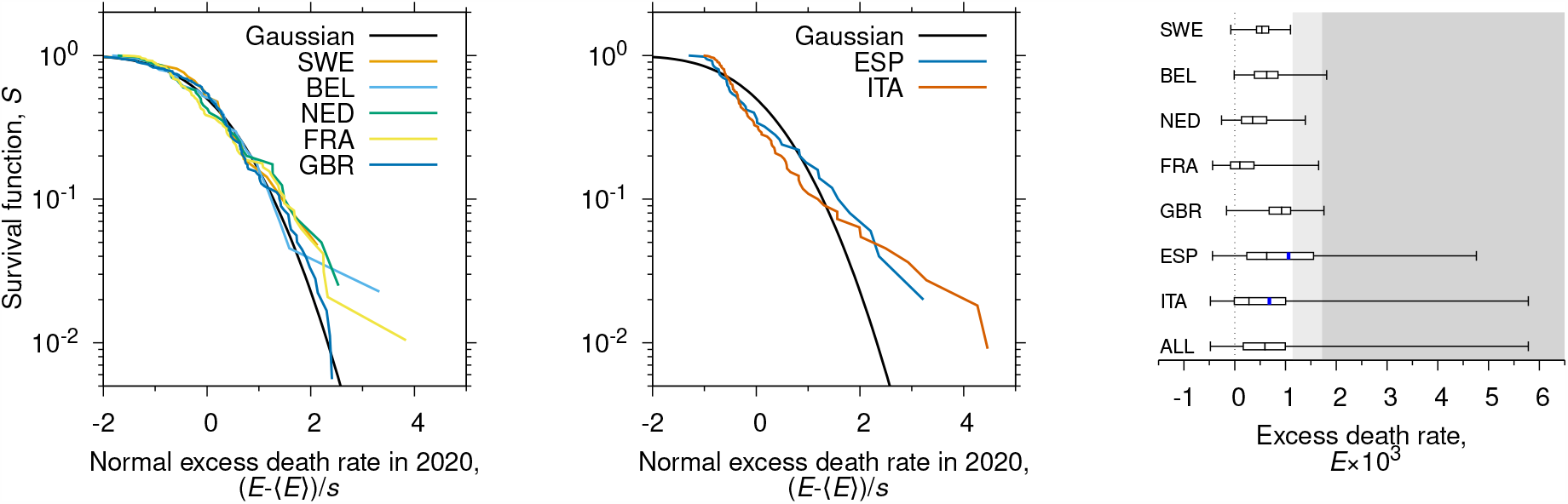
Empirical survival function versus the normal death rate excess in the spring of 2020 in Sweden, Belgium, Netherlands, France and United Kingdom (left); and in Spain and Italy (center). The Gaussian survival function (complementary error function) is noted in both pictures. Descriptive statistic values are listed in Table I. Canary Islands (ES7) and French overseas departments (FRY) were not included in the analysis. On the right a box-and-whisker plot of the distributions with blue marks highlighting the average *E* when it is different from the median (Spain and Italy). Background colors are explained in Figure 2.

## III. METHODS

### III.1. Accumulated excess death rates

Accumulated death rates —ratio of crude deaths to resident population— from W10 —week 10— to W21 in 2020 were computed for Netherlands, Belgium, France, and Spain; for Italy the interval run from W08 to W20; for United Kingdom from W12 to W22; for Sweden from W12 to W26. In 2020 W10 started on March 2 and W21 ended on May 24. Loosely speaking the intervals are representative of the spring of 2020. They were suggested from visual inspection of the death rate anomaly in 2020 and from the *z*− score anomalies observed in maps produced by EuroMoMo^5^.

A baseline or reference value can be obtained analyzing the accumulated death rate in the previous years. For that purpose a Poisson regression —`glm` function with option `family=“poisson”` in `R`— with a rate linearly varying with calendar year[7] was performed to get a predicted, baseline, reference death rate for 2020. The accumulated excess death rate of the interval —excess death rate hereafter— was then computed as the difference from the observed value in 2020 to the predicted baseline. Figure 5 Supplementary Material shows the results of this procedure for the largest anomalies recorded in every country.

### III.2. Statistics

The distribution of accumulated excess death rates {*E*_*i*_} in the spring of 2020 for territorial units will be described by its empirical survival function, the probability of finding an excess death rate larger or equal than a given value: *S* = *P* (*X*) ⩾ *x*).

An estimate for the survival function is obtained from a set of *N* observations *E*_*i*_ by ranking them in descending order of *E*_*i*_. The survival function is given by *S*_*k*_ = *k/N*, where *k* is the rank —the *k*th largest observation—. Notice that *S* must decrease with increasing *E*.

The KS distance 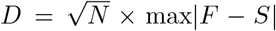, where *F* is the survival function of a continuous distribution like the Gaussian distribution, assesses whether the null hypothesis “empirical data originated from the tested distribution or *S* −*F* = 0” is or is not rejected at a given level of significance. For the standard level of significance *α* = 0.05 the KS distance equals to *D*_*c*_ = 1.36, irrespective of *N*. Notice that larger empirical samples need smaller absolute differences to overshot the critical threshold and reject the null.

## IV. RESULTS

Table I lists descriptive values for the excess death rate statistics: number of territorial units *N*, 2020 population *P*, population to number of territorial units and total excess death toll —computed as∑ _*i*_ *E*_*i*_ *× P*_*i*_—, followed by the median excess death rate *Q*_2_, the average excess death rate *⟨E⟩*, the standard deviation of excesses *s*, the ratio excess death toll to population or population weighted excess average *E*_*w*_, the largest excess death rate and its rank; and the KS distance to the Gaussian distribution. The null hypothesis is rejected for the Italian distribution and the combined distribution only. Swedish, Belgian, Dutch and British excess death rates find low KS distances.

**TABLE 1.** Descriptive statistic values level 3 of territorial units per country and all six combined. First, the number of territorial units, population numbers for 2020 and the ratio population to number of territorial units; then, excess death toll in the spring of 2020; followed by median excess death rate, averages excess death rate and standard deviation; the ratio excess death toll to population —population weighted average excess death rate—; the largest event in the country and its rank; and the KS distance to the Gaussian distribution. Values in blue ink highlight the rejection of the null hypothesis at the standard level of significance (*p* < 0.05). French overseas departments (FRY) and Canary Islands (ES7) did not enter in the analysis.

|  | SWE | NED | BEL | FRA | ITA | ESP | GBR | ALL |
| --- | --- | --- | --- | --- | --- | --- | --- | --- |
| Number of NUTS3 | 21<br>3.9 % | 40<br>7.4 % | 44<br>8.1 % | 96<br>17.8 % | 110<br>20.4 % | 50<br>9.3 % | 179<br>33.1 % | 540 |
| Population (2020) | 10 327 589<br>3.7 % | 17 407 585<br>6.2 % | 11 549 888<br>4.1 % | 67 098 824<br>24.1 % | 60 244 639<br>21.6 % | 45 092 671<br>16.2 % | 67 025 542<br>24.0 % | 278 746 738 |
| Pop per NUTS3 | 491 790 | 435 190 | 262 497 | 698 946 | 547 679 | 901 853 | 374 444 | 516 198 |
| Excess death toll | 6450<br>3.2 % | 8104<br>4.1 % | 7993<br>4.0 % | 19 399<br>9.8 % | 43 236<br>21.8 % | 48 938<br>24.6 % | 64 444<br>32.5 % | 198 564 |
| Median excess $Q_2$<br>($\times 10^6$) | 534 | 353 | 631 | 101 | 280 | 632 | 924 | 597 |
| Average excess $\langle E \rangle$<br>($\times 10^6$) | 546 | 407 | 636 | 188 | 682 | 1059 | 904 | 673 |
| Standard deviation $s$<br>($\times 10^6$) | 260 | 387 | 353 | 382 | 1143 | 1149 | 354 | 745 |
| Excess death to pop-<br>ulation ($\times 10^6$) | 625 | 466 | 692 | 289 | 718 | 1085 | 961 | 712 |
| Largest excess death<br>rate ($\times 10^6$)/rank | 1099/91 | 1390/53 | 1810/24 | 1653/30 | 5783/1 | 4761/3 | 1758/25 | |
| KS distance to Gaus-<br>sian distribution | 0.41 | 0.52 | 0.44 | 1.23 | 1.97 | 1.12 | 0.83 | 2.71 |

Figure 1 shows the empirical survival function *S* = *P* (*X*): ⩾*x*) of the excess death rate *E* during the spring of 2020 for Sweden, Belgium, Netherlands, France and United Kingdom (left), Spain and Italy (center); and a box-and-whisker plot (right). Background colors are later referenced to Figure 2. Note that *S* is presented in logarithmic scale, which allows a better visualization of the largest events. The empirical survival function accommodates to a Gaussian distribution in the left-most panel. The Gaussian survival function is the complementary error function and falls to zero as exp(− *x*^*2*^)*/x*. In contrast, the empirical distribution in Spain and in Italy (center) finds log *S* linear with *E*, an exponential distribution with little curvature. Average excesses differ signficantly from median excesses (see also Table I).

**FIG. 2.**
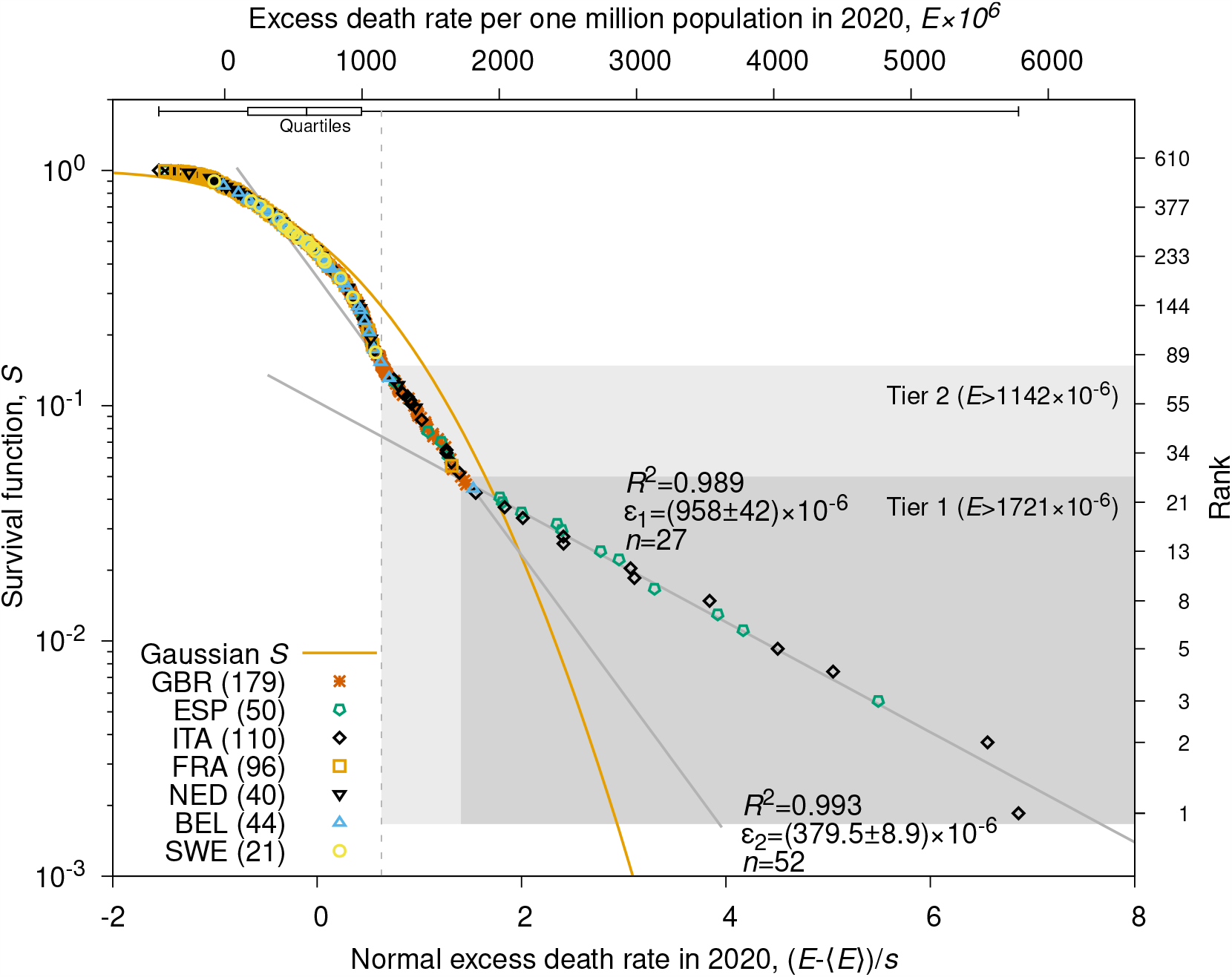
The empirical distribution of excess death rates in the *N* = 540 level 3 territorial units of Netherlands, Belgium, France, Spain, Italy and United Kingdom against the normal of the excess death rate score (*E* − *⟨E⟩*)*/s*. Different color and symbol according to country are used. The Gaussian survival function is noted by an orange line. Gray backgrounds display the first tier (dark, *k* < 27) and the second tier, *k* < 80). Bivariate analysis results for *S ∝* 2^−*E/ε*^ are annotated in the plot. The vertical broken line shows the position of the KS distance to the Gaussian distribution *D* = 2.71 (*p* < 10^−6^) which is computed as the vertical largest difference from the data points to the Gaussian line scaled by 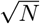.

The empirical survival function for the *N* = 540 territorial units for statistics is plotted against the excess death rate (top axis) and its normal score (bottom axis) in Figure 2. Quartile box and whisker are shown in the top of plot area. The location of the KS distance is also noted by a vertical broken line.

Three regimes can be identified. First, a rounding region in the highest ranked region (*k >* 80, 85 % of the distribution) not much different from what is expected for a standard Gaussian distribution.

Second, in the middle of the distribution curvature falls to zero and an exponential distribution of events log_2_ *S* = *a* + *b*_2_*E* or 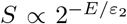 is noted (tier 2). The characteristic excess death rate that halves the empirical survival function is *ε*_2_ = 379.5(89) *×* 10^−6^.

Finally the largest events (tier 1, *k* < 27) find also an exponential distribution but the characteristic excess death rate is *ε*_1_ = 958(42) *×*10^−6^. The critical excess death rate is *E*_*c*_ = 1721*×* 10^−6^. The crossover of the last two regions marks the onset of a tail markedly heavier than the Gaussian tail with large events occurring at abnormally large frequencies. Tier 1 is noted by a dark gray background in the figure; tier 2, by a light gray background both in Figure 2 and in the right most panel of Figure 1.

The distribution of the *m* = 80 largest events and of the *N*− *m* = 460 smallest events is shown in Table 1 and Table 2 (Supplementary Material). The largest events occurred mostly in Great Britain (37), Italy (22) and Spain (16) although it must be noted that 32 % of Spanish territorial units and 38.2 % of Spanish population entered in this category, while in Great Britain that made 20.7 % and 24.2 %. Likewise 79.1 % (ESP) and 75.0 % (ITA) of the excess death toll occurred in this category but only a 34.1 % in United Kingdom. On the contrary, the whole Swedish population and the overwhelming majority of population and excess death toll in Netherlands, Belgium and France are found in territorial units belonging to the lowest tier of the distribution. Table 3 in Supplementary Material lists the 50 largest and the 50 smallest events.

## V. DISCUSSION

The analysis is conditioned by the distribution of level 3 territorial units within countries. While the main goal of territorial units for statistics is to provide homogeneous regions the outcome is far from being optimum. The average population per territorial unit ranges from 200*×* 10^3^ (Belgium) to 900 *×*10^3^ (Spain) (see Table I).

The countries here analyzed displayed the largest records of excess death rate in Europe during the spring 2020 (see Figures 1 to 4 in Supplementary Material). Therefore the exponential distribution of large events in the tier 1 (*k ⩽* 27; *E >* 1730 × 10^−6^; dark gray background in Figure 2) with characteristic death rate *ε*_1_ × 968 10^−6^ is a robust result since adding more countries to the analysis would only vertically shift *S* from *k/N* to *k/*(*N* + *n*), where *n* is the number of new territorial units, for these large events. Tier 1 is populated almost exclusively by Italian and Spanish territorial units (see Figure 2 and Table 1 in Supplementary Material). Tier 2 (27 < *k ⩽* 80; 1721 × 10^−6^ *> E* ⩾ 1142 × 10^−6^; light gray background in figure 2) is also a robust result: a visual inspection of Figure 4 in Supplementary Material suffices to understand that no other region outside those analyzed in this work should get an accumulated excess death rate larger than 1140 *×*10^−6^ (tier 2). In sharp contrast excess death rates outside tier 1 and 2 (*E* < 1142 *×* 10^−6^, *N* = 460) are normally distributed.

The results suggests that when the size of the outbreak was relatively low, its outcome —measured as the excess death rate— was affected by interaction with neighbouring regions and it was normally distributed. On the contrary, when the size of the outbreak was relatively high, specifically in tier 1, the outcome resulted from an isolated environment. In other words, once the disease set in a region, and once the size of infections, and community spread was above some threshold, and perhaps, once lock-down measures and social distancing came into play, flows of persons across boundaries played a lesser role and the outcome. As a result, the distribution turned into an exponential distribution, which appears in independently distributed events. In practical terms this occurred in Spain and Italy (see Figure 1).

## VI. CONCLUSION

Excess death rates in Europe during the 2020 spring outbreak of the illness COVID–19 were distributed exponentially above *E*_*c*_ *∼*1721 × 10^−6^ with with empirical survival function halving every *ε*_1_ = 958(42) × 10^−6^.

A second tier exponentially distributed tier of excess death rates is found for 1142 × 10^−6^ *⩽ E* < *E*_*c*_ with a characteristic halving excess death rate *ε*_2_ = 379(8) *×*10^−6^. On the contrary smaller events were distributed normally.

## Data Availability

All data were retrieved from Eurostat and the Spanish National Statistics Institute.

## ACKNOWLEDGMENTS

The author wishes to thank Eurostat and the National Statistics Institutes in Europe for releasing week crude deaths, and Mr. José María López-Cepero for his help with Eurostat database.

This work benefited from `GNU octave-5.2.0` (https://www.gnu.org/software/octave/); `R-3.4.4` (https://www.r-project.org/); `gnuplot-5.2.8` (http://www.gnuplot.info/); `GNU emacs-26.3` (https://www.gnu.org/software/emacs/) and AUCTEX(https://www.gnu.org/software/auctex/). This project started on September 12, 2020.

## Footnotes

1 filename: `demo r mweek3.tsv.gz` https://ec.europa.eu/eurostat/estat-navtree-portlet-prod/BulkDownloadListing?sort=1&downfile=data%2Fdemo_r_mweek3.tsv.gz (accessed September 2020)

2 filename: `demo r pjangr3` https://ec.europa.eu/eurostat/estat-navtree-portlet-prod/BulkDownloadListing?file=data/demo_r_pjangrp3.tsv.gz (accessed September 2020)

3 See https://ec.europa.eu/eurostat/tgm/table.do?tab=table&plugin=1&language=en&pcode=tps00001 (accessed September 2020)

4 See https://ine.es/jaxiT3/Tabla.htm?t=31304&L=1 (accessed September 2020)

5 See https://www.euromomo.eu/graphs-and-maps#map-of-z-scores

## References

[1] Janine Aaron, John Mullbauer, Charlie Giattino, and Hannah Ritchie, “A pandemic primer on excess mortality statistics and their comparability across countries,” (2020).

[2] Na Zhu, Dingyu Zhang, Wenling Wang, Xingwang Li, Bo Yang, Jingdong Song, Xiang Zhao, Baoying Huang, Weifeng Shi, Roujian Lu, Peihua Niu, Faxian Zhan, Xuejun Ma, Dayan Wang, Wenbo Xu, Guizhen Wu, George F. Gao, and Wenjie Tan, “A novel coronavirus from patients with pneumonia in China, 2019,” New England Journal of Medicine 382, 727–733 (2020).

[3] Annie Campbell and Edward Morgan, “Comparisons of all-cause mortality between European countries and regions : January to June 2020,” Office for National Statistics, 1–27 (2020).

[4] Aaron Clauset, Cosma R Shalizi, and MEJ Newman, “Power-law distributions in empirical data,” SIAM Review 51, 661–703 (2009).

[5] Pasquale Cirillo and Nassim Nicholas Taleb, “Tail risk of contagious diseases,” Nature Physics 16, 606–613 (2020), arXiv:2004.08658.

[6] Alvaro Corral, “Scientific comment on “Tail risk of contagious diseases”,” (2020), arXiv:2007.06876.

[7] Jose Maria Martin-Olalla, “Age and sex disaggregation of crude excess deaths during the 2020 spring COVID-19 outbreak in Spain,” medRxiv, 2020.08.06.20169326 (2020).

